# Primate-specific ZNF808 is essential for pancreatic development in humans

**DOI:** 10.1101/2021.08.23.21262262

**Authors:** Elisa De Franco, Nick D L Owens, Hossam Montaser, Matthew N Wakeling, Jonna Saarimäki-Vire, Hazem Ibrahim, Athina Triantou, Diego Balboa, Richard C Caswell, Matthew B Johnson, Sian Ellard, Caroline F Wright, Pancreatic Agenesis Gene Discovery Consortium, Sarah E Flanagan, Timo Otonkoski, Andrew T Hattersley, Michael Imbeault

**Author notes:** These authors contributed equally (Elisa De Franco, Nick D L Owens, Hossam Montaser). These authors contributed equally (Timo Otonkoski, Andrew T Hattersley, Michael Imbeault).

## Abstract

Identifying genes linked to extreme phenotypes in humans has the potential to highlight new biological processes fundamental for human development. Here we report the identification of homozygous loss of function variants in the primate-specific gene *ZNF808* as a cause of pancreatic agenesis. ZNF808 is a member of the KRAB zinc finger protein (KZFPs) family, a large and rapidly evolving group of epigenetic silencers that target transposable elements. We show that loss of *ZNF808 in vitro* results in aberrant activation of many transposable elements it normally represses during early pancreas development. This results in inappropriate specification of cell fate with induction of genes associated with liver endoderm and a loss of pancreatic identity. We show that ZNF808 and its transposable element targets play a critical role in cell fate specification during human pancreatic development. This is the first report of loss of a primate-specific gene causing a congenital developmental disease and highlights the essential role of ZNF808 for pancreatic development in humans.

## Introduction

Human genetic studies have provided key insights into differences in pancreatic development between human and mouse, highlighting multiple evolutionary conserved transcription factors with different dosage-dependent effects between the two species^1^. However, there are unexplained yet fundamental differences in the genetics, development, cell structure, and physiology of insulin-producing beta cells between human and rodent pancreas^2,3^. Identifying the genetic causes of rare congenital diseases, such as pancreatic agenesis, can highlight genes which are essential for organ development in humans. Here, we report the identification of the primate-specific gene *ZNF808* as a novel genetic cause of pancreatic agenesis, a condition resulting from defective fetal pancreatic development.

ZNF808 is a member of the KRAB zinc finger protein (KZFPs) family, the largest group of DNA binding factors in the human genome with around 350 genes. KZFPs primarily act as epigenetic silencers of transposable elements (TE) through establishment of heterochromatin-associated H3K9me3. The KZFP family is rapidly evolving, with new members found at most phylogenetic branches since its emergence at the dawn of tetrapods. KZFPs co-evolve with TEs in an arms race aimed at suppressing their replication^4^. A large survey of KZFPs and their binding sites has revealed that many are evolutionary conserved at various levels (for example within primates or within mammals) along with remnants of their TE targets. These conserved KZFPs are hypothesized to control chromatin accessibility of regulatory platforms derived from conserved TEs, playing a role in the evolutionary rewiring of gene regulatory networks^5^.

## Results

### Identification of *ZNF808* homozygous variants as a cause of pancreatic agenesis

To identify novel genetic causes of pancreatic agenesis, we performed exome sequencing in two unrelated affected individuals and their parents. Both patients were born from a consanguineous union and were diagnosed with pancreatic agenesis, defined as insulin-dependent diabetes diagnosed in the first 6 months of life (neonatal diabetes) and exocrine pancreatic insufficiency^6^. The two patients were found to be homozygous for 11 and 12 rare coding non-synonymous variants respectively (Supplementary tables 1 and 2). The only gene found to harbour homozygous variants in both patients was *ZNF808*, with a deletion of exons 4 and 5 identified in proband 1 and a nonsense variant (p.Leu213*) detected in proband 2.

To replicate this finding, we investigated the presence of rare *ZNF808* biallelic variants in 232 additional patients diagnosed with neonatal diabetes before the age of 6 months without an identified pathogenic variant in the 30 known aetiological genes^7^. Homozygous loss of function *ZNF808* variants (6 nonsense, 3 frameshifts, and 1 whole-gene deletion) were identified in an additional 10 unrelated individuals (Figure 1A, Supplementary table 3). Family testing confirmed that all unaffected parents were heterozygous carriers for the identified *ZNF808* loss of function variants. In one family a sibling diagnosed with diabetes in age-range 39-52 weeks, had the same homozygous *ZNF808* mutation as the proband (Sup. table 3). All frameshift and nonsense variants affect residues within the last exon of the gene containing all 23 zinc finger domains (which comprises 93% of the protein) and are predicted to result in a truncated protein lacking from 3 to 23 of the 23 functional zinc finger domains. Two patients (probands 2 and 3) were homozygous for large deletions predicted to result in no *ZNF808* mRNA.

**Figure 1.**
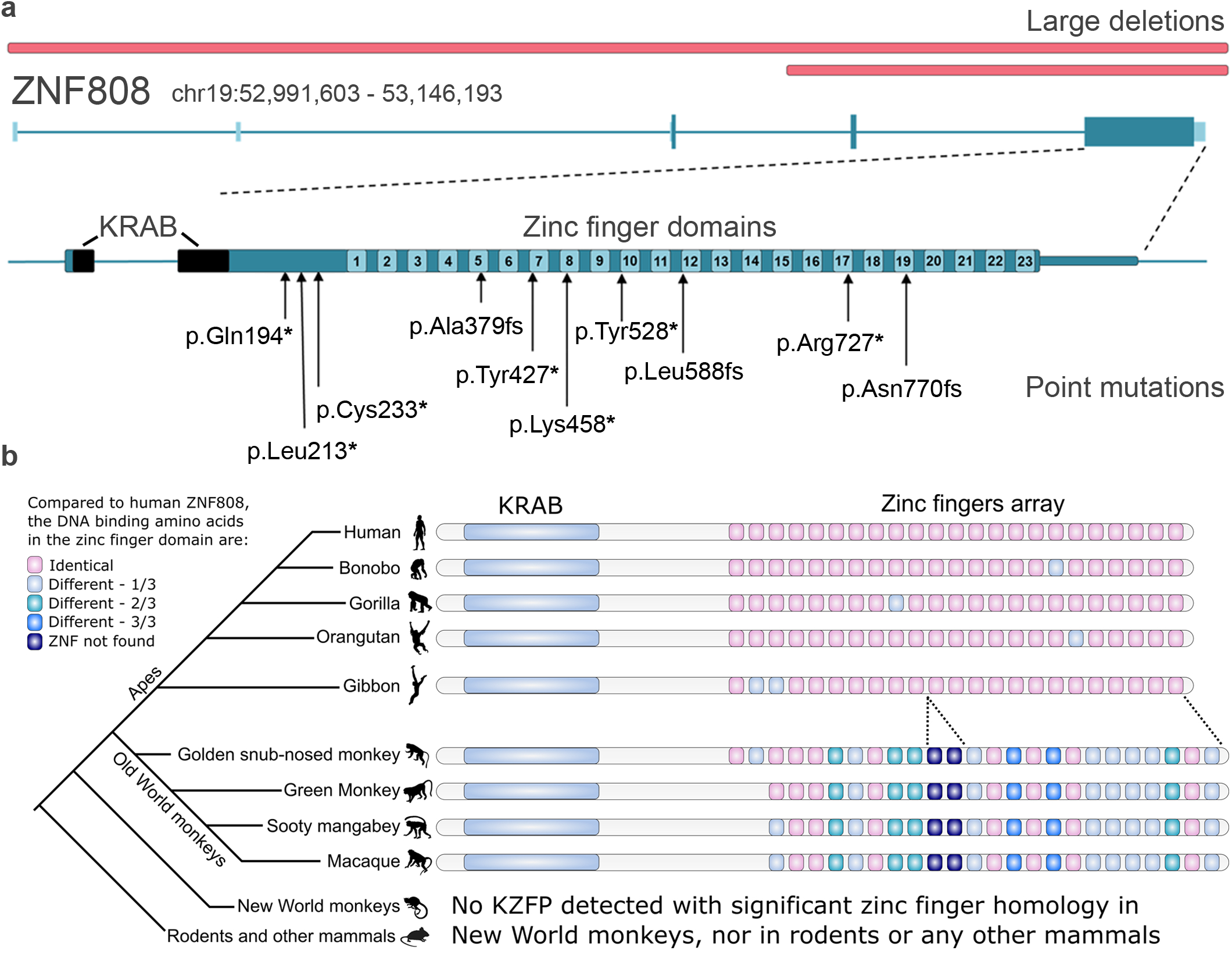
Genetics, discovery and evolutionary conservation of ZNF808. **A. Schematic representation of the *ZNF808* gene and of the identified mutations**. Deletions identified at this locus in patients are highlighted in red above the gene cartoon while various loss of function mutations we have identified are represented below. Protein domains such as KRAB and zinc finger are annotated at the gene level. **B. Reconstructed phylogeny of ZNF808 using a zinc finger signature approach**. The three amino acids directly binding DNA were used to track evolution of the zinc finger array using data from ^5^. Zinc finger domains are color-coded according to the number of mutations in each triplet compared with the human version. Significant events of loss or gain of zinc fingers are also represented. No significant homology with any zinc finger array was detected in New World monkeys nor in any other mammals. Silhouettes of representative species were downloaded from http://phylopic.org.

Despite ZNF808 being widely expressed in human tissues, the patients’ phenotype is restricted to the pancreas, with diabetes onset in the neonatal period (median age at diagnosis 12 weeks, IQR 3-21) and low birth weight (median −2.98 SD, IQR −3.56 to −2.47) which results from markedly reduced insulin-mediated growth in utero (Supplementary table 3). Pancreatic agenesis was diagnosed in 5/5 patients for whom exocrine pancreatic function through fecal elastase measurement was available and was suspected in two further cases who presented clinical symptoms of malabsorption, suggesting a likely developmental defect affecting both the exocrine and endocrine pancreatic functions. No additional extra-pancreatic features were consistently observed in the cohort. The extremely low birth weight, early age at diagnosis of diabetes, and exocrine pancreatic insufficiency all suggest a role of ZNF808 in early pancreatic development. This is supported by previous human embryo transcriptomic studies showing that ZNF808 is expressed in the embryonic pancreas^8^.

### ZNF808 and its transposable element targets are primate-specific

The evolutionary origin of ZNF808 can be traced to a common ancestor of Old World monkeys and Apes, with a very high level of conservation in Apes (Figure 1B). Between Old World monkeys and humans, the absence of the first two zinc fingers, two additional zinc fingers in the middle of the array, and a number of amino acid differences in DNA binding residues of most zinc fingers can be observed. Importantly, no KZFP with a similar array of zinc fingers could be found in New World monkeys nor any other mammals. Genome-wide patterns of ZNF808 show that it primarily targets the long terminal repeat (LTR) of endogenous retroviruses classified as MER11 (A/B/C) elements (Figures 2A). Analysis of mammalian genomes shows that these transposons also originate in Old World monkeys and Apes (Figure 2B). Last, we found that MER11 elements are also targeted by other primate-specific KZFPs (ZNF433, ZNF440, ZNF468 and ZNF525) (Figure 2C). Therefore, ZNF808 and its targeted MER11 elements are exclusively found in primates, making it the first primate-specific genetic cause identified for a congenital developmental disease (Supplementary Figure 1).

**Figure 2.**
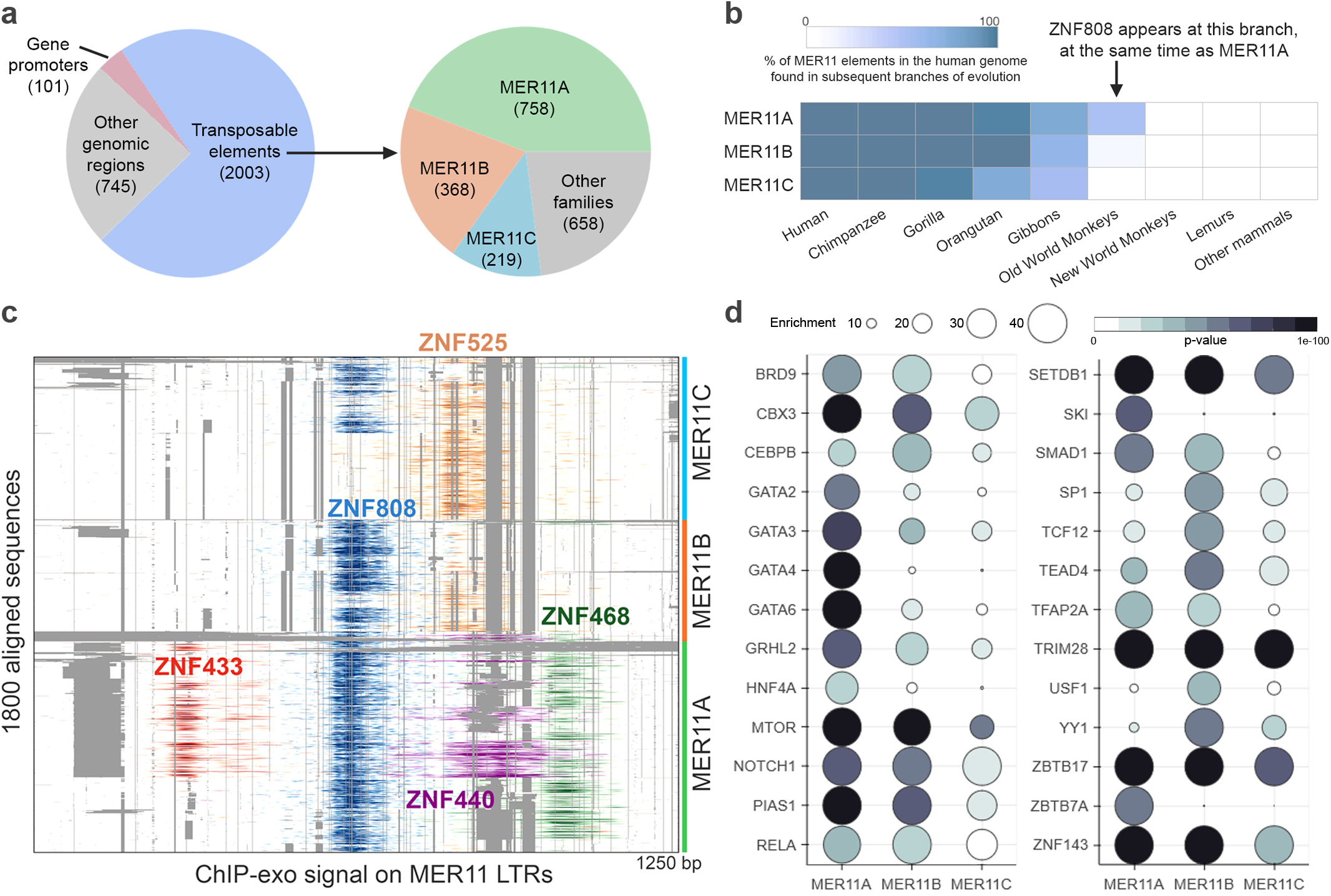
ZNF808 is the main transcriptional repressor of domesticated primate-specific endogenous retroviruses. **A. ZNF808 binds primarily MER11 transposable elements**. Analysis of ZNF808 ChIP-seq data^5^ reveals that it primarily intersects with transposable elements, although a minority of binding sites are found on gene promoters and other genomic regions. Further analysis of transposons shows that ZNF808 binds primarily elements of the MER11 family – MER11A, MER11B and MER11C. **B. MER11 transposable elements are primate specific**. The origin of each individual MER11 element in the human genome was traced using a comparative multiple alignment strategy with the genome of 100 other species. The age of each element was determined as being corresponding to the farthest phylogenetic branch where we could find a similar copy at a syntenic locus. The graph shows the cumulative sum of elements at each phylogenetic branch, starting from mammals. Here it is clear that MER11 elements originated in Old World monkeys and replicated successfully for approximately 15 million years before losing transposition capabilities. **C. ZNF808 is the main KZFP binding MER11 elements, but other primate-specific KZFPs bind select subsets**. Overlay of signal from published KZFPs ChIP-seq on a multiple alignment of MER11A, MER11B and MER11C elements reveal that ZNF808 binds strongly in the centre of these elements. Four other KZFPs can be found on smaller subsets of elements, sometimes redundantly with ZNF808, but with a clear pattern of semi-exclusive binding with ZNF525. All KZFPs binding MER11 elements represented here were found to be primate-specific. **D. MER11 elements bound by ZNF808 are enriched for DNA binding factors and their interactors**. The main DNA binding factors along with secondary interactors found to be enriched (p-value < 1e-30 and enrichment > 30) in various MER11 subfamilies are represented here. The colour of each bubble is scaled with p-value and the radius with enrichment. If multiple experiments are found to be enriched for any given factor, only the most significant value is shown. The full ChIP-Atlas dataset contains 15217 experiments of 880 different DNA binding factors and their interactors at the time of analysis.

### MER11 elements are enriched in transcription factor binding sites involved in early embryonic development

Correlative studies have previously suggested that MER11 elements might be a source of regulatory potential in multiple tissues^9–11^. Our analysis of publicly available ChIP-seq data shows that various subfamilies of MER11 elements bound by ZNF808 show enriched occupancy for a set of 26 DNA binding factors (Figure 2D), including some transcription factors involved in early embryonic development. Importantly, three of these (GATA4, GATA6, and HNF4A) have been previously described as monogenic causes of pancreatic agenesis and diabetes^6,13–15^. This evidence suggests that the transposable elements silenced by ZNF808 might be domesticated regulatory platforms and could play a functional role in primate pancreas development.

### Loss of ZNF808 reveals active epigenetic potential of MER11 elements during pancreatic differentiation

To characterize the molecular events triggered by *ZNF808* loss in an *in vitro* model of pancreas development, we functionally inactivated *ZNF808* in H1 human embryonic stem cells using CRISPR, obtaining a clone with in-frame deletions resulting in the loss of 8 zinc fingers (*ZNF808*^*del/del*^) (Supplementary figure 2A). Using this clone, we assayed epigenetic changes resulting from the functional impairment of ZNF808, both in stem cells and at key steps of differentiation towards beta cells (Figure 3A and Supplementary figure 2C)^16^. Using ChIP-seq, we quantified the genome-wide presence of H3K9me3, a hallmark of heterochromatin that is induced by KZFPs. H3K9me3 was detected in around half the known binding sites of ZNF808, with the vast majority being transposons (mostly MER11 elements, data not shown). We observed loss of H3K9me3 at a subset of these TEs in the *ZNF808*^*del/del*^ mutant at early stages of differentiation, ranging from 64% of sites in ES cells to 31% in primitive gut tube (Figure 3B). However, the loss was not complete even when considering subfamilies of elements. MER11A were most affected (497 sites), followed by MER11B (254 sites) and MER11C (123 sites), approximately following the prevalence of ZNF808 binding in these families (Figure 3B). The presence of other primate-specific KZFPs redundantly binding MER11 elements as mentioned above could explain the incomplete loss of H3K9me3.

**Figure 3.**
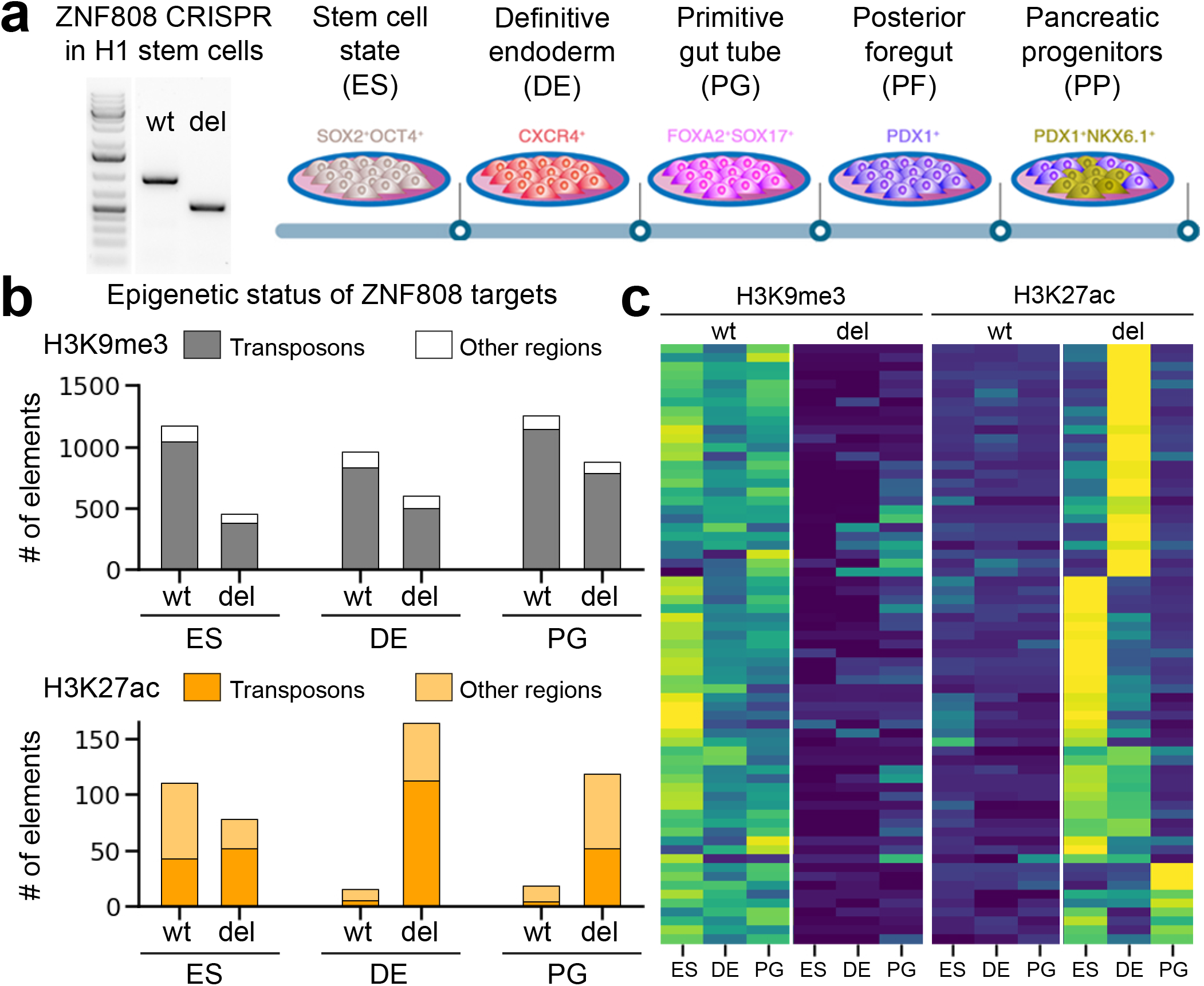
Loss of ZNF808 during pancreas differentiation leads to the aberrant epigenetic reactivation of MER11 elements. **A. ZNF808 functional impairment using CRISPR**. Left - H1 stem cells exposed to Cas9 and two guide RNAs targeting the zinc finger array were used to produce *ZNF808*^*del/del*^. This plot shows the PCR amplification of the region targeted in both wildtype H1 stem cells and the isolated clone with a successful deletion, showing the expected shorter size confirming deletion of 8 zinc fingers in the ZNF808 array. Right - an overview of the multi-step differentiation protocol used in this study for both epigenetic and transcriptomic analysis of differences induced by *ZNF808*^*del/del*^. **B. ZNF808 is important for the maintenance of epigenetic repression on transposons**. H3K9me3 ChIP-seq peaks intersecting with ZNF808 binding sites reveal that a majority of sites are covered in heterochromatin-associated H3K9me3 and are transposable elements. A large proportion of these peaks lose H3K9me3 in *ZNF808*^*del/del*^ clones at various stages of differentiation. On the other hand, analysis of shows that many sites gain H3K27ac, especially at the early stages of differentiation – this is more evenly split between transposon and other regions bound by ZNF808. **C. Heatmap showing a selected subset of epigenetically dynamic targets of ZNF808**. Clustering all ZNF808 peaks showing either H3K9me3 or H3K27ac allowed us to identify a subset showing loss of H3K9me3 followed by gain of H3K27ac in at least one stage of differentiation. Normalized ChIP-seq signal minus input is shown – colour scale ranges from +2 (yellow) to –1 (blue) on a z-score scale.

We next surveyed if the loss of silencing revealed regulatory potential at ZNF808 binding sites. In embryonic stem cells (ES), a number of sites were already positive for H3K27ac, but no significant change could be detected in the *ZNF808*^*del/del*^ clone in terms of total positive H3K27ac sites. However, during differentiation, we observed the emergence of H3K27ac epigenetic marks in 149 ZNF808 binding sites at the definitive endoderm stage and 100 sites at the primitive gut tube stage (Figure 3B). We could identify a subset of 67 MER11 elements where loss of H3K9me3 was followed by gain of H3K27ac during differentiation in the *ZNF808*^*del/del*^ clone (Figure 3C). Altogether, these results show that ZNF808 is normally silencing MER11 elements during differentiation toward pancreatic cells and that the loss of ZNF808 triggers epigenetic activity associated with transcription from a subset of them, suggesting that these could function as alternative promoters or enhancers.

### Upon ZNF808 loss, activated MER11 elements lead to upregulation of nearby genes at early and late stages of pancreatic differentiation

We next quantified transcriptomic changes induced in the *ZNF808*^*del/del*^ during our differentiation protocol. We found that the number of genes significantly dysregulated (FDR < 0.05, absolute fold change > 1.25) varies from 473 at the ES stage to 565 at definitive endoderm (DE), followed by a marked peak of 3858 and 3670 differentially regulated genes at the primitive gut tube (PG) and posterior foregut (PF) stages, respectively, reducing to 683 at the pancreatic progenitor (PP) stage (Figure 4A). We investigated to what extent the dysregulation of MER11 elements might be responsible for these gene expression changes by assessing the enrichment of genes changing expression in proximity to ZNF808 binding sites. Genes activated at the ES and DE stages were enriched in proximity to MER11 elements at enhancer but not promoter distances, and we find no particular proximity enrichment for genes repressed (Figure 4B). We find clear examples of robustly upregulated genes in close proximity to the set of MER11 elements that lose H3K9me3 and gain H3K27ac at early stages (Figure 4C). The proximity enrichments diminish at the PG and PF stages but do partially recover at the PP stage suggestive of roles for MER11 elements throughout pancreatic cell differentiation. The loss of proximity enrichment at the PG stage concomitant with large increase in dysregulated genes, suggests that these genes are involved in expression programmes downstream of MER11 activation.

**Figure 4.**
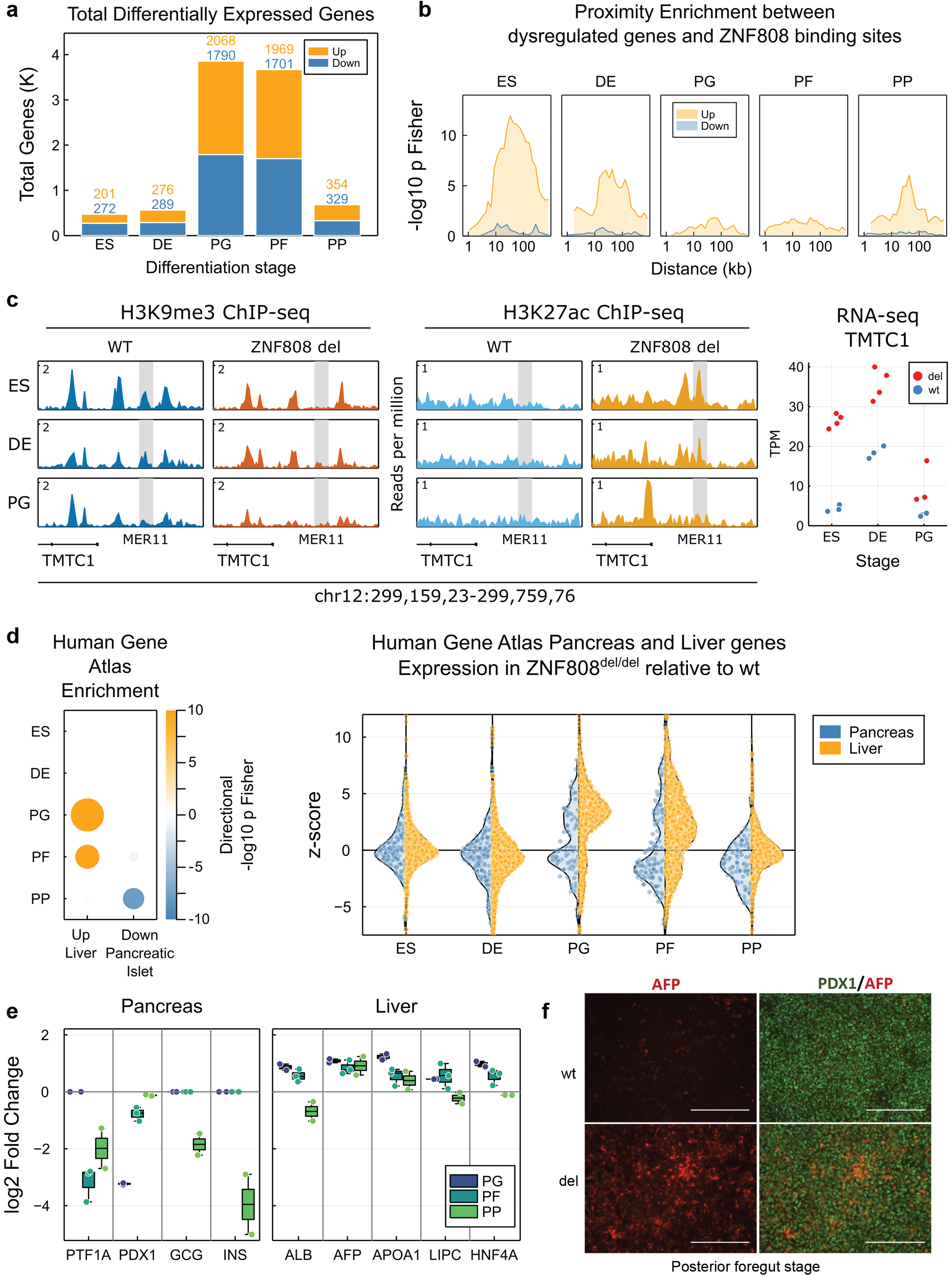
Loss of ZNF808 during pancreas differentiation leads to a repression of pancreatic lineages and activation of liver lineages, driven by gene expression changes in proximity to MER11 elements. **A. Loss of ZNF808 leads to perturbed gene expression throughput pancreatic differentiation**. Bar chart showing total genes up/activated and down/repressed with FDR < 0.05 and |FC| > 1.25 for each stage of pancreatic differentiation. **B. Dysregulated genes are found in proximity to ZNF808 binding sites at the ES and DE stages**. Proximity enrichment (-log10 Fisher exact p-value, right tail) showing an excess of dysregulated genes in proximity to ZNF808 peaks as a function of distance between genes and binding sites for genes up/activated (orange) and down/repressed (blue). Enrichment peaks between 10kb-100kb consistent with ZNF808 repressing distal gene enhancers. **C. Example locus showing loss of repression and activation of transposon, with activation of adjacent gene**. Locus shows H3K9me3 and H3K27ac data in reads per million upstream of the TMTC1 promoter, in which H3K9me3 marked MER11 element in wild type is lost at all stages in ZNF808^del/del^ concomitant with gain H3K27ac at the ES cell stage. Gene expression for TMTC1 shown (right) in transcripts per million (TPM), showing robust upregulation at ES, DE and PG stages. **D. Genes activated in *ZNF808***^***del/del***^ **are enriched in liver specific expression and genes repressed are enriched in pancreas specific expression. Left -**Enrichment using Enrichr and Human Gene Atlas gene set for Liver and Pancreas terms for genes dysregulated at each stage of pancreatic differentiation, area of dot proportional to –log10 p Fisher and colour indicates up/activated and down/repressed. **Right -** Trajectory of all genes in Human Gene Atlas Pancreas (left, blue) and Liver (right, orange) annotations over pancreatic differentiation stages, mean z-score of ZNF808^del/del^ relative to wild type, each dot represents a single gene. **E. Repression and activation of selected respective pancreatic and liver lineage genes**. Boxplots and dotplots showing log2 Fold change *of ZNF808*^*del/del*^ relative to mean wild type expression for pancreas and liver markers at PG, PF and PP stages. Each gene is significantly dysregulated with FDR < 0.05, |FC|>1.25 for at least one stage. **F. Immunostaining of AFP and PDX1 at PF stage**. Confirmation of RNA-seq results by immunostaining, showing activation of AFP in ZNF808^del/del^ in PDX1 positive cells (scale bar = 100 um).

### *ZNF808* inactivation leads to dysregulation of genes defining pancreatic and liver identity during pancreatic differentiation

To understand how this gene expression dysregulation impacts pancreatic development we performed gene set enrichment analysis using Enrichr^17^. We first associated genes activated or repressed in *ZNF808*^*del/del*^ at each stage of the *in vitro* pancreatic differentiation protocol to genes with specific tissue expression in the Human Gene Atlas. We observed a large enrichment of genes known to be expressed in liver at the PG stage, that precedes a downregulation of genes specifically expressed in the pancreas PF and PP stages (Figure 4D - left). Assessing the trajectory of pancreatic and liver programmes, we see the downregulation of pancreatic genes and the upregulation of liver genes beginning from the PG stage and continuing until the PP stage (Figure 4D – right). Indeed, transcription factors necessary for pancreatic development, such as PTF1A and PDX1, show reduced expression at PG and PF stages, which is followed by reduced expression of the endocrine markers insulin and glucagon at the PP stage (Figure 4F). In parallel with these events, we see activation of early markers of liver differentiation (including AFP, ALB and HNF4A, the latter of which has roles in specifying pancreas and liver) at the PG and PF stages (Figure 4E), as confirmed by immunostaining for AFP (Figure 4F). In further support of this hypothesis, we examined the relationship between our data and a survey of the transcriptome of human embryo dorsal pancreas buds and hepatic cords obtained by laser dissection^8^. At the PG stage, we found a comprehensive enrichment with 266 of the 2068 genes activated in *ZNF808*^*del/del*^ having increased expression in hepatic cords and 181 of 1790 genes repressed in in *ZNF808*^*del/del*^ having decreased expression in dorsal pancreas buds (Fishers Exact p < 10^−56^, Odds Ratio 4.02 for genes activated; p < 10^−25^, OR 2.82 for repressed). Taken together this data suggests that ZNF808 acts to balance the differentiation of endoderm progenitors to liver or pancreas lineages.

## Discussion

In this manuscript we report the identification of *ZNF808* as critical for human pancreatic development, the first example of a primate-specific gene implicated in the aetiology of a congenital developmental disease in humans. This finding offers novel insights into the marked differences between human and rodent pancreas and further confirms the crucial importance of gene discovery efforts in individuals with extreme phenotypes, as the role of ZNF808 could not have been identified through mouse studies.

This discovery also provides one clear example where a KZFP and its TE target have been co-domesticated to rewire gene regulatory networks. Although the mammalian-conserved ZFP568 was found to play a critical role in regulating IGF2 in early embryogenesis, no transposable element was found at the causal binding sites^18^. While the biological function of most KZFPs remains to be elucidated, the implication of primate-specific ZNF808 and MER11 transposable elements in such an important evolutionary process as pancreas development suggests that KZFPs have the potential to be collectively involved in other important aspects of human biology. Our work confirms that the regulatory potential contained in domesticated transposable elements is only starting to be revealed. There are 150 KZFPs shared between human and most mammals, the vast majority of which with no known biological function, highlighting the need to further investigate KZFPs targeting ancient remnants of transposons.

Our results reveal that loss of ZNF808 results in the aberrant induction of a gene expression programme normally found in liver endoderm during *in vitro* differentiation toward pancreatic lineages. This suggests that a cell fate diversion from pancreas to liver lineages may be the pathogenic mechanism that results in neonatal diabetes and exocrine insufficiency in patients with biallelic loss of function mutations in *ZNF808*, with ZNF808 acting to balance these two lineages. Our results imply that both KZFPs and their MER11 elements targets have been co-domesticated to regulate multiple stages of endoderm derived organogenesis, with genes dysregulated found in proximity to MER11 elements at early and late stages of differentiation. This also suggests that MER11 element are active later or elsewhere in development and that ZNF808 is necessary to ensure they are repressed at early stages of pancreas development. To fully untangle the network of regulation established by ZNF808 will require accounting for the many KZFPs that bind subsets of MER11 elements., These together could form a complex combinatorial system of regulation, with select subsets of MER11 elements allowed to be active in particular cellular contexts depending on the combination of KZFPs and transcription factors present. Indeed, all five KZFPs identified to bind MER11 elements have dynamic patterns of expression, including during pancreatic development (data not shown).

Finally, this work has highlighted the role of a primate-specific KZFP, *ZNF808*, and its TE targets in the regulation of human beta cell development, providing important insights into human development and diabetes pathogenesis, which will support efforts aimed at providing cell-based therapies for people with diabetes.

**Materials and methods** in Supplementary Appendix.

## Data Availability

All data will be made available on publication in a peer reviewed publication. All disease-causing variants identified by next generation sequencing will be uploaded onto the DECIPHER database (https://decipher.sanger.ac.uk/). All variants identified through genome sequencing will be submitted as a .vcf file with allele frequencies to the EMBL-EBI European Genome-Phenome Archive (EGA) (https://www.ebi.ac.uk/ega/home).

All epigenomic and transcriptomic data from in vitro pancreatic differentiation will be deposited in NCBI Gene Expression Omnibus (GEO, https://www.ncbi.nlm.nih.gov/geo/).

https://www.ncbi.nlm.nih.gov/geo/query/acc.cgi?acc=GSE78099

https://www.ebi.ac.uk/arrayexpress/experiments/E-MTAB-3259/

## Acknowledgements

We are grateful to the patients and their families for taking part in our gene discovery study. We also thank Solja Eurola and Jarkko Ustinov for expert technical assistance.

