## Supplementary appendix for "Primate-specific ZNF808 is essential for pancreatic development in humans"

**Supplementary note**

**Members of the Pancreatic Agenesis Gene Discovery Consortium**

Wafaa Laimon[1], Saman S Hassan[2], Mohamed A Abdullah[3], Anders Fritzberg[4], Emma Wakeling[5], Nisha Nathwani[6], Nancy Elbarbary[7], Amany Osman[8], Hessa Alkandari[9-10,] Abeer alTararwa[10], Abdelhadi Habeb[11], Abdulmoein Eid Al-Agha[12], Ihab Abdulhamed Ahmed[13], Majida Noori Nasaif[14], Ala Ustyol[15]

**Institutions:**

[1]Pediatric Endocrinology and Diabetes Unit, Department of Pediatrics, Mansoura Faculty of Medicine, Mansoura University Children's Hospital, Mansoura University, Mansoura, Egypt.
[2]Gaafar Ibn Auf Pediatric Tertiary Hospital, Khartoum, Sudan
[3]Faculty of Medicine, University of Khartoum, Khartoum, Sudan
[4]Region of Dalarna – Child and adolescent medicine Falun Hospital, Falun, Sweden
[5]North East Thames Regional Genetic Service, Great Ormond Street Hospital for Children NHS Foundation Trust, London, UK
[6]Bedfordshire Hospitals NHS Foundation Trust, Luton and Dunstable Hospital NHS Foundation Trust, Luton, UK
[7]Diabetes Unit, Department of Pediatrics, Faculty of Medicine, Ain Shams University, Cairo, Egypt
[8]Imperial College London Diabetes Centre, Al Ain,United Arab Emirates
[9]Department of Population Health, Dasman Diabetes Institute, Kuwait
[10]Department of Paediatrics, Farwaniya Hospital, Kuwait
[11]Paediatric department, Prince Mohammed bin Abdulaziz Hospital, Madinah, KSA
[12]Paediatric department, King Abdulaziz University Hospital, Jeddah, Saudi Arabia
[13]Pediatric Endocrinology Unit, Zagazig University Hospital, Zagazig, Egypt
[14]Dr. Soliman Fakeeh Medical Center, Al-Basateen, Jeddah, Saudi Arabia
[15]Department of Pediatrics, University of Health Sciences Haseki Training and Research Hospital, İstanbul, Turkey.

**Supplemental Methods**

**Subjects.**

Individuals with neonatal diabetes diagnosed before the age of 6 months were recruited by their clinicians for molecular genetic analysis in the Exeter Molecular Genetics Laboratory. The study was conducted in accordance with the Declaration of Helsinki, and all subjects or their parents/guardian gave informed consent for genetic testing.

**Genetic analysis.**

Exonic sequences were enriched from genomic DNA using Agilent's SureSelect Human All Exon kit (version 4) and then sequenced on an Illumina HiSeq 2000 sequencer using 100- bp paired-end reads. The sequencing data were analyzed using an approach based on the GATK best practice guidelines. GATK HaplotypeCaller was used to identify variants that were annotated using Alamut batch version 1.8 (human reference genome assembly hg19), and variants that failed the QD2 VCF filter or had less than 5 reads supporting the variant allele were excluded. Copy number variants were called by SavvyCNV, which uses read depth to judge copy number states. SavvyVcfHomozygosity was used to identify large (>3 Mb) homozygous regions in the exome sequencing data (https://github.com/rdemolgen/SavvySuite).

Replication studies were performed in a cohort of 232 patients diagnosed with diabetes before age 6 months in whom the known genetic causes of neonatal diabetes had been excluded. Patients were analyzed using a targeted next-generation sequencing assay, which includes baits for known neonatal diabetes genes and additional candidate genes followed up from gene discovery, such as *ZNF808*, or by independent genome sequencing analysis. Variant confirmation and co-segregation in family members were performed by Sanger sequencing (primers available on request).

**Cell culture and in vitro differentiation of human embryonic stem cells.**

Human embryonic stem cells (hESC) (WA01/H1 line, Wicell) were cultured on Matrigel-coated plates (BD Biosciences) in Essential 8 (E8) medium (Life technologies, A1517001) and passaged using EDTA. Differentiation experiments were carried out on confluent plates of stem cells, following a seven-stage protocol as previously described[1].

**Genome editing.**

To create an in vitro model for studying the role of ZNF808 in pancreatic development, guide RNAs targeting the zinc fingers domain of the fifth exon of ZNF808 for deletion were designed using Benchling (https://benchling.com). gRNAs with the highest quality score and lowest off-targets score were selected and purchased, alongside the RNP components (HiFi Cas9 protein, crRNA and tracrRNA), from Integrated DNA Technologies (IDT) and utilized according to the manufacturer's recommended protocol. Two million cells were electroporated with the RNP complex using Neon Transfection system (Thermo Fisher, 1100 V, 20ms, two pulses), and plated on Matrigel-coated plates in E8 medium containing 10 µM ROCK inhibitor overnight. Afterwards, cells were single-cell sorted, expanded, and screened for the desired deletion using PCR. Positive clones were validated by Sanger sequencing at Eurofins Genomics and the sequences were aligned using Geneious Prime 2020.1.1. Additionally, the KO clones were characterized for pluripotency, chromosomal integrity, and the top 3 off-target hits predicted by the online tool CRISPOR[2] were checked and no off-target indels were found.

**Formaldehyde crosslinking.**

To fix the cells for ChIP-seq samples preparation, cells were incubated with TrypLE for 5-10 min at 37°C and gently homogenized with the pipette, then pooled in a 15 ml Falcon containing warm DMEM. Afterwards, cells were spun down at 250 g at room temperature for 3 min, then resuspended in DMEM at a concentration of 5 million cells per mL and incubated with 333 mM fresh 16% methanol-free formaldehyde at room temperature for precisely 10 min. Formaldehyde was quenched using 250 mM Tris pH 8.0 for another 10 min at room temperature, then cells were spun down at 250g at 4°C for 5 min. Cell pellet was resuspended gently in PBS, aliquoted into 1.5 mL eppendorf tubes, and spun down at 250g at 4°C for 5 min. Supernatant was removed and samples were stored at -80°C till further processing.

**Flow cytometry analysis.**

hESC-derived cells were dissociated into single cells by incubation with TrypLE for 10 min at 37°C and resuspended in cold FBS/PBS (5% v/v). For surface marker staining of CXCR4, 1 million cells were incubated with the directly conjugated antibody CD184/CXCR4 APC at a final dilution of 1:10 for 30 min at room temperature. For intracellular markers, 1 million cells were first fixed in 350 µl of Cytofix/Cytoprem Buffer (BD, No. 554722) for 20 min at 4°C, then washed twice with BD Perm/Wash Buffer Solution (BD, No. 554723). Cell pellet was resuspended in 80 µl FBS/BD Prem/Wash buffer (4% v/v) and incubated with the corresponding directly conjugated antibody at a final dilution of 1:80 overnight at 4°C. After incubation with the antibody, cells were washed twice and analyzed using FACSCalibur cytometer (BD Bioscience) and FlowJo software v9 (Tree Star Inc.). Details of the used antibodies are listed in supplementary table 4.

**Immunocytochemistry.**

For adherent cultures, cells were fixed in 4% PFA for 15 min at RT, permeabilized with 0.5% triton-X100 in PBS, then blocked with UltraV block (ThermoFisher) for 10 min and incubated with primary antibodies diluted in 0.1% Tween in PBS overnight at 4°C. After incubation, cells were washed twice with PBS and incubated with corresponding secondary antibodies diluted in 0.1% Tween in PBS for 1 h at room temperature. Details of the used antibodies are listed in supplementary table 4.

**Chromatin immunoprecipitation (ChIP)**

The following steps were performed with ice-cold samples and buffers containing a protease inhibitor cocktail (cOmplete ULTRA Tablets EDTA-free, Roche). In 1.5mL DNA LoBind tubes, 4 million fixed cells were resuspended in 1 mL lysis buffer 1 (50mM HEPES-KOH pH 7.4, 140mM NaCl, 1mM EDTA, 0.5mM EGTA,

10% Glycerol, 0.5% NP40, 0.25% Triton X-100, Proteinase Inhibitor 1x) incubated at 4$^{o}$C on a rotating wheel at 10 RPM for 10 min. Cells were then centrifuged at 1700 g for 5 min at 4$^{o}$C. Supernatant was discarded and pellets were resuspended in 1mL lysis buffer 2 (10mM Tris HCl pH 8.0, 200mM NaCl, 1mM EDTA, 0.5mM EGTA, Proteinase Inhibitor 1x) and incubated at 4$^{o}$C on a rotating wheel at 10 RPM for 10min. After centrifugation at 1700 g for 5 min at 4$^{o}$C, supernatant was discarded and pellets were washed with 500 uL SDS shearing buffer (1mM Tris HCl pH 8.0, 1mM EDTA, 0.15% SDS, Proteinase Inhibitor 1x), without disturbing the pellets followed by centrifugation at 1700 g for 5 min. Washing was repeated two times and pellets were resuspended in 1 mL SDS shearing buffer and transferred into Covaris milliTUBE 1 ml AFA Fiber. Chromatin was sheared on a Covaris E220 for 6 min at 5% duty cycle, 140W, 200 cycles. The sheared chromatin was then transferred into 1.5mL DNA LoBind tubes and centrifuged at 10000 g for 5min at 4$^{o}$C. Supernatant was then used immediately for immunoprecipitation. Chromatin quality control was performed on Bioanalyzer 2100 (Agilent) to verify that most fragments were ranging between 200 and 600 bp.

For H3K9me3 IP, chromatin corresponding to 1 million cells was put in a new 1.5 mL DNA LoBind tube and topped to 900 uL with SDS shearing buffer. For H3K27ac IP, chromatin corresponding to 3 million cells was put in a new 1.5mL DNA LoBind tube and topped to 900uL total with SDS shearing buffer. IP conditions were adjusted to 150 mM NaCl and 1% Triton final. 1 uL of anti-H3K9me3 antibody (Active Motif, 39161) or 5 ug of anti-H3K27ac antibody (Active Motif, 39685) was added. The IP was then incubated on rotating wheel at 10RPM at 4$^{o}$C overnight. The next day, Dynabeads Protein G (5uL for the H3K9me3 IP or 25uL for the H3K27ac) were put on magnet and supernatant was removed. Beads were then resuspended in the full volume of the IP and incubated for 2 hours on a rotating wheel at 10 RPM at 4$^{o}$C.

For H3K9me3, low salt washing buffer (10mM Tris HCl pH 8.0, 150mM NaCl, 1mM EDTA, 1% Triton X100, 0.15%SDS, 1mM PMSF) and high salt washing buffer (10mM Tris HCl pH 8.0, 500mM NaCl, 1mM EDTA, 1% Triton X100, 0.15%SDS, 1mM PMSF) were used.

For H3K27ac, low salt washing buffer (20mM Tris-HCl pH 8, 150mM NaCl, 2mM EDTA, 1% Triton X-100, 0.1% SDS, 1mM PMSF) and high salt washing buffer (20mM TrisHCl pH 8, 500mM NaCl, 2mM EDTA, 1% Triton X-100, 0.1% SDS, 1mM PMSF) were used.

All washes took place while IPs and buffers were ice cold. PMSF was always added in the buffers immediately before each wash. The IPs were placed on a magnetic rack and supernatant was discarded. Beads were resuspended in low salt washing buffer and transferred into a clean DNA LoBind tube. Beads were then placed on a magnetic rack; supernatant was removed, and beads were resuspended in low salt washing buffer. The mixture was placed again on a magnetic rack, supernatant was discarded, and beads were washed with high salt washing buffer. Once more, samples were placed on the magnetic rack, supernatant was removed and beads were resuspended in LiCl buffer (10mM Tris HCl pH8.0, 1mM EDTA, 0.5mM EGTA, 250mM LiCl, 1%NP40, 1%NaDOC, 1mM PMSF). The mixture was placed on the magnetic rack, supernatant was removed, and beads were washed with 10mM Tris HCL pH 8.0 and transferred to a clean DNA LoBind tube. Finally, with the samples on the magnetic rack the supernatant was completely removed, and beads were resuspended in Elution buffer (10mM Tris-HCl pH 8.0, 1mM EDTA, 1% SDS and 150mM NaCl).

RNase A was added to the elution buffer at a final concentration of 0.5ug/uL and samples were incubated at 37$^{o}$C for 1h in a shaking incubator at 1100 RPM. Subsequently proteinase K was added at a concentration of 400 ng/µl and chromatin was decrosslinked at 65$^{o}$C overnight. The supernatant was collected and purified using serapure beads before library preparation.

*Quality control of the IP*

To control for the efficiency of the IP, we used qPCR with primers targeting negative and positive region in the genome for the histone marks H3K9me3 (K9_Neg_GAPDH_Fw: CACCGTCAAGGCTGAGAACG, K9_Neg_GAPDH_Rv: ATACCCAAGGGAGCCACACC, K9_Pos_ZNF555_Fw: CAATTGGCCCATATCTTTACG, K9_Pos_ZNF555_Rv: CATGTTCTCGAAAGCAAGCA) and H3K27ac (K27_Neg_Fw: TACACATCAGCCATTCTTACAG, K27_Neg_Rv: GCTAATCTTAAATTCCACTCTCCC, K27_Pos_EIF4A2_Fw: GGGGAAAGCGAGGTTTAACT, K27_Pos_EIF4A2_Rv: TTACAGGGTCGCTGGAAATC) respectively.

*Library preparation*

For the library preparation we used NEBNext® Ultra™ II DNA Library Prep Kit for Illumina®. The protocol provided by the manufacturer was followed. The library was quantified with qpcr using KAPA SYBR® FAST and the set of primers itru7_101_01 and itru5_01_A. After quantification the library was amplified and double indexed using primers:

itru7_101_01: CAA GCA GAA GAC GGC ATA CGA GAT GGT AAC GTG TGA CTG GAG TTC A*G

itru7_101_02: CAA GCA GAA GAC GGC ATA CGA GAT CAA CAC AGG TGA CTG GAG TTC A*G

itru7_101_03: CAA GCA GAA GAC GGC ATA CGA GAT ACA CCT CAG TGA CTG GAG TTC A*G

itru5_01_A: AAT GAT ACG GCG ACC ACC GAG ATC TAC ACA CCG ACA AAC ACT CTT TCC CTA* C

itru5_01_B: AAT GAT ACG GCG ACC ACC GAG ATC TAC ACA GTG GCA AAC ACT CTT TCC CTA* C

itru5_01_C: AAT GAT ACG GCG ACC ACC GAG ATC TAC ACC ACA GAC TAC ACT CTT TCC CTA* C

itru5_01_D: AAT GAT ACG GCG ACC ACC GAG ATC TAC ACC GAC ACT TAC ACT CTT TCC CTA* C

itru5_01_E: AAT GAT ACG GCG ACC ACC GAG ATC TAC ACG ACT TGT GAC ACT CTT TCC CTA* C

itru5_01_F: AAT GAT ACG GCG ACC ACC GAG ATC TAC ACG TGA GAC TAC ACT CTT TCC CTA* C

Amplified libraries were double size selected using home-made serapure beads to enrich for fragments between 200 and 600bp.

Adapters and indexed primers design, resuspension and annealing were as previously described[3]. Adapter sequences were iTrusR2-stubRCp: /5Phos/GAT CGG AAG AGC ACA CGT CTG AAC TCC AGT CA*C, iTrusR1-stub: ACA CTC TTT CCC TAC ACG ACG CTC TTC CGA TC*T

After amplification library was controlled to confirm that the IP remained efficient using qPCR with primers targeting negative and positive region in the genome for the histone marks H3K9me3 and H3K27ac respectively, as mentioned above. Libraries were sent for 150 basepairs paired end sequencing at Novogene.

*ChIP-seq analysis*

Reads from each library were mapped on hg19 using Bowtie2 version 2.4.4[4] with the 'very-sensitive-local' setting. Mapped reads were compressed in BAM files and indexed using SAMtools version 1.18[5]. We used MACS version 1.4.3[6] to call peaks for H3K9me3 and H3K27ac datasets, using the corresponding input datasets as background controls.

Enrichment of transcription factors was made using the Enrichment Analysis tool from ChIP-Atlas (https://chip-atlas.org[7]) using MER11 elements of distinct subfamilies (MER11A, MER11B, MER11B) intersecting with ZNF808, with hg19 and a score 50 as a target threshold and 100 iterations of random permutations.

**Gene expression analysis by RNA-seq**

Stranded, poly-A selected RNA-seq libraries were prepared and sequenced (paired-end 150bp reads) by Novogene Co Ltd.

Stranded paired end 150bp RNA-seq reads were aligned to the hg19 genome using STAR[8] and quantified against Gencode v36 release liftover to hg19 by RSEM[9] using the RSEM-STAR pipeline, with additional options "--seed 1618 --calc-pme --calc-ci --estimate-rspd --paired-end". RSEM estimated read counts per sample were rounded for use with DESeq2[10]. We perform differential expression analysis for each stage wt vs del for each for all genes with at least 10 raw counts in all replicates of one condition. Gene expression varies significantly over the differentiation timecourse with subsets of genes only expressed at early or late stages, therefore when testing each stage we supply DESeq2 with samples from adjacent stages for information sharing in estimating dispersions. We consider genes with FC > 1.25 and FDR < 0.05 differentially expressed.

To determine enrichments of differentially expressed genes in proximity to clusters of epigenetic response in ZNF808 loss, we used the package ProximityEnrichment.jl (https://github.com/owensnick/ProximityEnrichment.jl). Specifically, we calculate Hypergeometric right tail p-values for the association of differentially expressed genes within *x* bp of ZNF808 bound region cluster against a background of genes for x in [1, 1e+6]. For proximity enrichment heatmaps, we take the maximal enrichment from this interval.

To determine gene set enrichments for sets of differentially expressed genes, we use the Enrichr API[11], with the package (https://github.com/owensnick/Enrichr.jl) to recover enrichments for Human_Gene_Atlas gene set. To assess the intersection between our data and the laser capture of human embryo hepatic cords and dorsal pancreas[12], we took the set of genes detected in each stage of our data and hepatic cords vs dorsal panc, and calculated the association between direction of dysregulation in our data with the direction of genes differentially expressed genes in the hepatic cords vs dorsal panc comparison with FDR < 0.05.

### Supplemental tables

#### Supplementary table 1– Homozygous rare (MAF<0.001) variants with at least 5 supporting reads identified in Patient 1

| Gene | Coding Effect | Allele depth | gNomenclature | cNomenclature | pNomenclature | GnomAD allele frequency |
|---|---|---|---|---|---|---|
| *PRMT6* | missense | 0,129 | Chr1(GRCh37):g.107599878G>C | NM_018137.2:c.541G>C | NM_018137.2:p.Gly181Arg | 0 |
| ***ZNF808*** | **stop gain** | **0,177** | **Chr19(GRCh37):g.53056806del** | **NM_001039886.3:c.637del** | **NM_001039886.3:p.Leu213*** | 0 |
| *SLC10A2* | missense | 0,97 | Chr13(GRCh37):g.103701740A>G | NM_000452.2:c.818T>C | NM_000452.2:p.Ile273Thr | 0.000004 |
| *PPARGC1A* | missense | 0,92 | Chr4(GRCh37):g.23815971G>A | NM_001330751.1:c.1150C>T | NM_001330751.1:p.Arg384Trp | 0.000018 |
| *FAM71E2* | missense | 0,43 | Chr19(GRCh37):g.55871059C>T | NM_001145402.1:c.1177G>A | NM_001145402.1:p.Gly393Arg | 0.00002 |
| *DCLRE1B* | missense | 0,76 | Chr1(GRCh37):g.114454395T>G | NM_001319946.1:c.803T>G | NM_001319946.1:p.Leu268Trp | 0.000024 |
| *DLGAP4* | missense | 0,174 | Chr20(GRCh37):g.35060206G>A | NM_014902.5:c.86G>A | NM_014902.5:p.Arg29His | 0.000097 |
| *SPAG4* | missense | 0,33 | Chr20(GRCh37):g.34205087G>A | NM_001317931.1:c.103G>A | NM_001317931.1:p.Glu35Lys | 0.000109 |
| *SSC5D* | missense | 0,37 | Chr19(GRCh37):g.56012117G>A | NM_001144950.1:c.2563G>A | NM_001144950.1:p.Ala855Thr | 0.000192 |
| *HERC1* | missense | 0,52 | Chr15(GRCh37):g.64048910G>A | NM_003922.3:c.1259C>T | NM_003922.3:p.Thr420Met | 0.000307 |
| *SNAPC5* | missense | 0,19 | Chr15(GRCh37):g.66790035T>A | NM_001329613.1:c.35A>T | NM_001329613.1:p.Glu12Val | 0.000747 |

**Supplementary table 2 – Homozygous rare (MAF<0.001) variants with at least 5 supporting reads identified in Patient 2**

| Gene | Coding Effect | Allele depth | gNomenclature | cNomenclature | pNomenclature | GnomAD allele frequency |
|---|---|---|---|---|---|---|
| *FAM135B* | missense | 0,16 | Chr8(GRCh37):g.139190807A>G | NM_015912.3:c.1000T>C | NM_015912.3:p.Tyr334His | 0.000012 |
| *KIDINS220* | missense | 0,13 | Chr2(GRCh37):g.8897901T>C | NM_001348745.1:c.3031A>G | NM_001348745.1:p.Ser1011Gly | 0.000061 |
| *EPAS1* | missense | 0,35 | Chr2(GRCh37):g.46607460G>A | NM_001430.4:c.1649G>A | NM_001430.4:p.Arg550Gln | 0.000065 |
| *EPPK1* | missense | 0,85 | LRG_880:g.14550G>A | NM_031308.3:c.4339G>A | NM_031308.3:p.Ala1447Thr | 0.00007 |
| *ZNF701* | CNV deletion | 0,0 | Chr19(GRCh37):g.53057128_53100968del | null | NM_001172655.1:null | 0 |
| ***ZNF808*** | **CNV deletion** | **0,0** | **Chr19(GRCh37):g.53057128_53100968del** | **null** | **NM_001039886.3:null** | **0** |
| *ARHGEF10* | missense | 0,68 | Chr8(GRCh37):g.1817407T>A<br>LRG_234:g.50259T>A | NM_001308152.1:c.673T>A | NM_001308152.1:p.Ser225Thr | 0.000122 |
| *LILRB1* | missense | 0,76 | Chr19(GRCh37):g.55148276G>A | NM_001081637.2:c.1906G>A | NM_001081637.2:p.Glu636Lys | 0.000138 |
| *TNFRSF9* | missense | 2,0 | Chr1(GRCh37):g.7993222G>A | NM_001561.5:c.679C>T | NM_001561.5:p.Pro227Ser | 0.000149 |
| *CROCC* | missense | 1,15 | Chr1(GRCh37):g.17296838G>A | NM_014675.3:c.5542G>A | NM_014675.3:p.Gly1848Arg | 0.000333 |
| *TTLL12* | missense | 0,20 | Chr22(GRCh37):g.43576914C>T | NM_015140.3:c.380G>A | NM_015140.3:p.Arg127His | 0.000432 |
| *AGPAT5* | missense | 0,56 | Chr8(GRCh37):g.6588258G>T | NM_018361.3:c.316G>T | NM_018361.3:p.Ala106Ser | 0.00096 |

### Supplemental table 3. Clinical features of patients with ZNF808 biallelic mutations

(to reduce potential indirect identifiers data provided is limited, birth weight given to nearest 200g, and age of diagnosis given as a range)

| ID | Sex | Known Related Parents | Mutation (all homozygous) | Birth weight (g) | Diabetes | Age at diagnosis of diabetes | Exocrine pancreatic insufficiency | Additional features |
|---|---|---|---|---|---|---|---|---|
| 1 | Male | No | p.Leu213* | 1400 | Yes | 0-13 wks | Yes (biochemically confirmed) | None |
| 2 | Female | Yes | Deletion of exons 4-5 | 1200 | Yes | 0-13 wks | Yes (biochemically confirmed) | None |
| 3 | Male | Yes | Whole gene deletion | 2000 | Yes | 14-26 wks | NA | None |
| 4 | Female | No | p.Gln194* | 1200 | Yes | 14-26 wks | Likely (clinical signs of malabsorption) | None |
| 5a | Male | Yes | p.Cys233* | 2400 | Yes | 14-26 wks | NA | None |
| 5b | Female | Yes | p.Cys233* | 2000 | Yes | 39-52 wks | NA | None |
| 6 | Female | Yes | p.Ala379Valfs*157 | 1600 | Yes | 0-13 wks | NA | None |
| 7 | Male | Yes | p.Tyr427* | 2600 | Yes | 0-13 wks | NA | None |
| 8 | Female | No | p.Lys458* | 1400 | Yes | 0-13 wks | Yes (biochemically confirmed) | Anemia |
| 9 | Male | No | p.Tyr528* | 2000 | Yes | 0-13 wks | Yes (biochemically confirmed) | None |
| 10 | Male | Yes | p.Leu588Profs*118 | 1600 | Yes | 0-13 wks | NA | None |
| 11 | Male | Yes | p.Arg727* | 1800 | Yes | 14-26 wks | Yes (biochemically confirmed) | None |
| 12 | Male | Yes | p.Asn770Ilefs*98 | 2000 | Yes | 14-26 wks | Likely (clinical signs of malabsorption) | None |

**Supplementary table 4** - **Antibodies used for flow cytometry (FC) and immunocytochemistry (ICC)**

| **Antibody** | **Supplier** | **Use** |
|---|---|---|
| Mouse Anti-CD184 (CXCR4) Monoclonal Antibody, PhycoErythrin Conjugated,Clone 12G5 | BD Biosciences Cat# 555974; RRID:AB_396267 | FC (1:10) |
| Mouse IgG2a, kappa Isotype Control, PhycoErythrin Conjugated, Clone G155-178 | BD Biosciences Cat# 563023 | FC (1:10) |
| Mouse Anti-PDX1 Phycoerythrin Conjugated | BD Biosciences Cat# 562161; RRID:AB_10893589 | FC (1:80) |
| Mouse Anti-NKX6-1 Alexa Fluor 647 Conjugated | BD Biosciences Cat# 563338 | FC (1:80) |
| Rabbit IgG Isotype Control (Alexa Fluor 647 Conjugate) | Cell Signaling Technology Cat# 3452S; RRID:AB_10695811 | FC (1:80) |
| Goat anti-PDX1 | R and D Systems Cat# AF2419; RRID:AB_355257 | ICC (1:250) |
| Mouse anti-NKX6-1 | DSHB Cat# F55A10; RRID:AB_532378 | ICC (1:250) |
| Rabbit anti-CHGA | Dako Cat# A0564 | ICC (1:500) |
| Guinea pig anti-INS | Dako Cat# A0564; RRID:AB_10013624 | ICC (1:500) |
| Rabbit anti-SOX9 | Millipore Cat# AB5535; RRID:AB_2239761R | ICC (1:500) |
| Sheep anti-NEUROG3 | R and D Systems Cat# AF3444; RRID:AB_2149527 | ICC (1:500) |
| Rabbit anti-AlphaFetoProtein (AFP) | Dako Cat# A0008 | ICC (1:350) |

**Supplemental figures**

**Supplementary Figure 1. ZNF808 is the first primate-specific gene reported to cause a congenital developmental disease.** Homology scores (including 1-to-1 and 1-to-many orthologues) for every protein coding gene in the human genome were downloaded from Ensembl Biomart (10th February 2020) using Ensembl 98 for 26 primates and 70 non-primate mammals. The Target%ID was extracted for each gene based on the human transcript with the maximum identity. The difference between the maximum homology across all primates versus the maximum across all non-primates was then calculated for each gene. Scores for individual human genes were binned into centiles and plotted from the largest difference between primates and non-primates (i.e. least well conserved between primates and non-primates) to the smallest difference (i.e. most well conserved). Genes were further grouped into: non-disease causing (i.e. not present in OMIM-morbid; black), disease-causing (i.e. present in OMIM-morbid; green), genetic causes of developmental disorders (i.e. present on DDG2P; red), and the location of ZNF808 is highlighted (blue dotted line). Manual check for mammalians orthologues and disease caused was carried out for the 10 OMIM morbid genes (green) with higher scores than ZNF808, showing that all of them either had a mouse orthologue (*KIAA1549* MGI:2669829, *TNFRSF10B* MGI:1341090; *LOR* MGI:96816, *HMGA2* MGI:101761, *CFH* MGI:88385, *NDUFA11* MGI:1917125, *GP6* MGI:1889810, *FLG2* MGI:3645678, *RPS24* MGI:98147) or did not cause a congenital developmental disease (*NLRP7* MGI:3041206, disease caused: Recurrent hydatidiform mole).

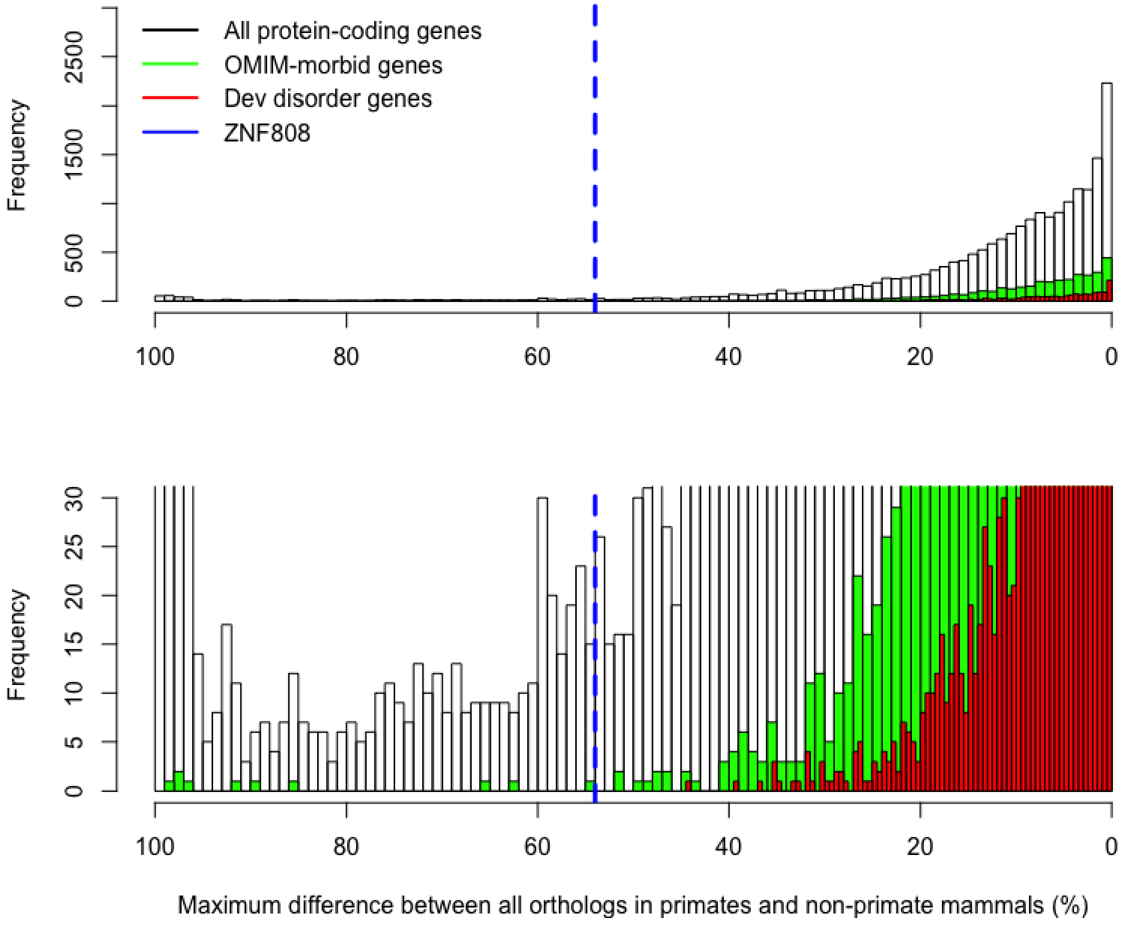

**Supplementary Figure 2. Generation and characterization of the *ZNF808*$^{del/del}$ cells.** A: Schematic presentation of the *ZNF808*$^{del/del}$ strategy using CRISPR/Cas9. The positions of the targeted regions are marked with red arrowheads. B: Sanger sequencing of the *ZNF808*$^{del/del}$ clone confirming the 401-nucleotide deletion. C: Flow cytometry analysis at different stages during in vitro differentiation: CXCR4 at Definitive endoderm (DE); PDX1+/NKX6.1+ at St4. D: Immunohistochemistry analysis of S4 adherent cells for PDX1, NKX6.1, chromogranin A (CHGA), Insulin (INS), SOX9 and NGN3 (Scale bar = 100 um)

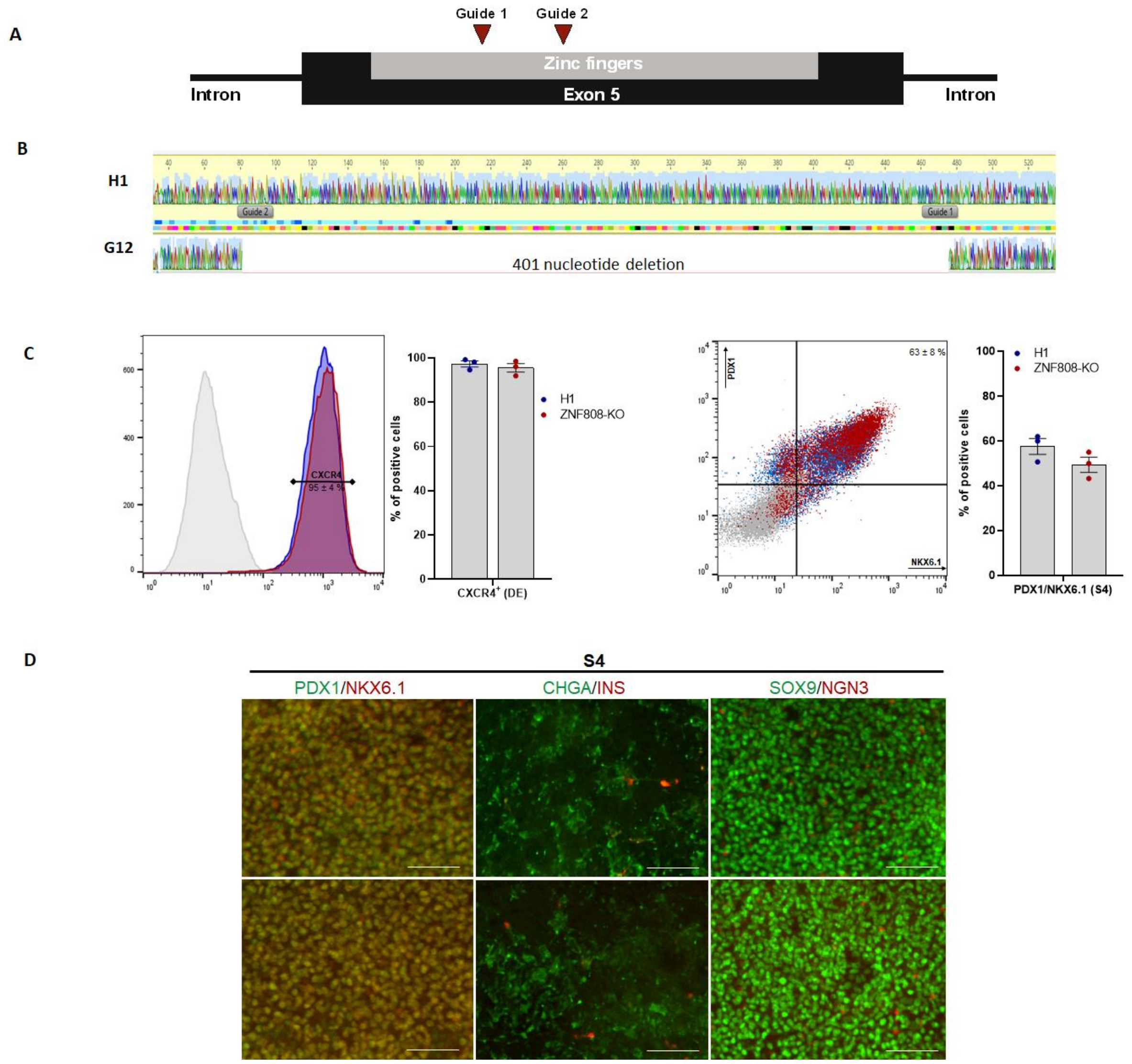
